# Segregation of children into small groups for in-person learning during the COVID-19 pandemic

**DOI:** 10.1101/2021.08.17.21261993

**Authors:** Luis Manuel Muñoz-Nava, Marcos Nahmad

## Abstract

The COVID-19 pandemic affected in-person learning worldwide due to fears that schools could contribute to the propagation of the virus within their communities. Using computational modeling, we compare the reopening of schools with mitigation measures with a strategy in which schoolchildren are segregated into small isolated groups or “bubbles,” where children physically interact without restrictions while receiving remote instruction from their teachers. This strategy is robust to common perturbations and is more flexible and stable than reopening of schools. Our modeling results and a real-world implementation of a bubbles program in an elementary school in Mexico City support that this strategy is an effective transition alternative, especially in communities with low vaccination rates or where operational costs associated to safely reopening schools cannot be afforded.

## Introduction

The COVID-19 pandemic affected more than 1.4 billion students worldwide (*1*). In many communities, school buildings have remained at least partially closed for more than a year and this situation may persist for several additional months while local vaccination efforts race skepticism, availability, and the surge of new variants of the virus (*2, 3*). Although the effects of the COVID-19 disease are generally mild in most schoolchildren, they could spread the disease to more vulnerable people, and a small percentage of children could experience serious complications such as multi-systemic immunological disorders (*4, 5*). Previous studies investigated how schools contribute to the propagation of the disease within their communities and concluded that, when mitigation measures are in place, schools are not hotspots for virus propagation (*6–10*). However, outbreaks may occur if prevention, testing, and isolation measures are not adequately implemented (*11, 12*). On the other hand, school closures are expected to have long-term effects on children education, social skills, and psychological behavior (*13–15*), and may impact negatively on parenting and family income (*16*). After balancing risks and benefits, public health experts are recommending the immediate reopening of schools (*17*) and are issuing operational and mitigation strategies to guide schools and local governments (*18*). Some key recommendations include: reducing class sizes, testing and contact tracing of positive cases, mask wearing, maintaining social distancing, appropriate ventilation in classrooms, and offering alternative online options for high-risk families (*17, 19, 20*). However, the implementation of adequate mitigation measures necessary to safely reopening school buildings faces several challenges: Can all schools afford the financial, logistic, and training costs to appropriately implement these measures? How would these measures affect the quality and equity of education? Given the high rate of asymptomatic or mild symptomatic cases in children, will the actions taken upon a detection of a positive case in a parent, a teacher, or a student be sufficient to contain a school-wide spread of the disease without constantly and widely interrupting in-person activities? How would schools, districts, and governments deal with parents and teacher unions’ resistance to in-person instruction?

Other than reopening of schools, alternatives for in-person learning exist and have been implemented empirically in several countries. Learning “pods,” “capsules,” or “bubbles” are groups of a few children that their parents voluntarily set-up for in-person instruction either from one of the parents or an external tutor. Informal learning bubbles were very popular and operated broadly in many countries in 2020. In the UK, the Department of Education asked schools to keep children in unmixed groups or bubbles, although later they removed this recommendation (*20*). Modeling studies on social segregation into groups controls for a rapid increase of COVID-19 infections (*21, 22*), but the effectiveness of segregating school communities into bubbles in mitigating the propagation of the COVID-19 disease and its impact on the operation of in-person learning programs has not been studied in detail. Moreover, it is unclear how such a strategy performs under different conditions, such as varying the size of the bubble or allowing children within a bubble not to obey social distancing or facemask wearing.

## Computational model

To investigate the effectiveness of segregating schoolchildren into bubbles, we took a computational modeling approach (see Materials and Methods for additional details). We modeled a school using a network of interacting nodes (representing children, parents, and/or teachers) interconnected by edges (representing social interactions within a family or bubble; Fig. 1A). In addition, all adults in the network (but not children) are connected to a master node (star symbol in Fig. 1A) encompassing other out-of-school interactions in the society (e.g., work or friends). At each time step, each node can be in one of the following states of the disease (Fig. 1B): Susceptible (S), representing that the individual has not contracted COVID-19; Infected (I), representing an individual that acquired the disease (I_1_ is the first day of infected and transitions in this state for four additional days, I_2_-I_5_); at day 6 after becoming infected, the individual either becomes Symptomatic (C) or remains Asymptomatic (A) for the following 9 days (C_i_ or A_i_ with i=1-9, respectively); after 14 days of becoming infected, the individual automatically transitions to a Recovered (R) state, which we assume is no longer affected by the disease (*i*.*e*., we assume that for the duration of our simulations, people can get the disease only once). The dynamics of the network are governed by a transition probability matrix, which takes into account the current state and the interactions of each node with its neighbors (directly connected through edges in the network; Fig. 1B and Materials and Methods). When an individual (child or adult) transitions to state C_1_, the edges of all bubbles in which the children of that family participate are removed for 14 days and then they are reinstated, except for those children in which any family member becomes symptomatic within these 14 days; in such a case, the 14-day isolation time is restated for that family (Fig. 1C). In our simulations, all nodes begin in the S state and network dynamics evolve for 90 days. COVID-19 infection probabilities were obtained from previous studies (Tables S1-S3 and Materials and Methods).

**Fig. 1.**
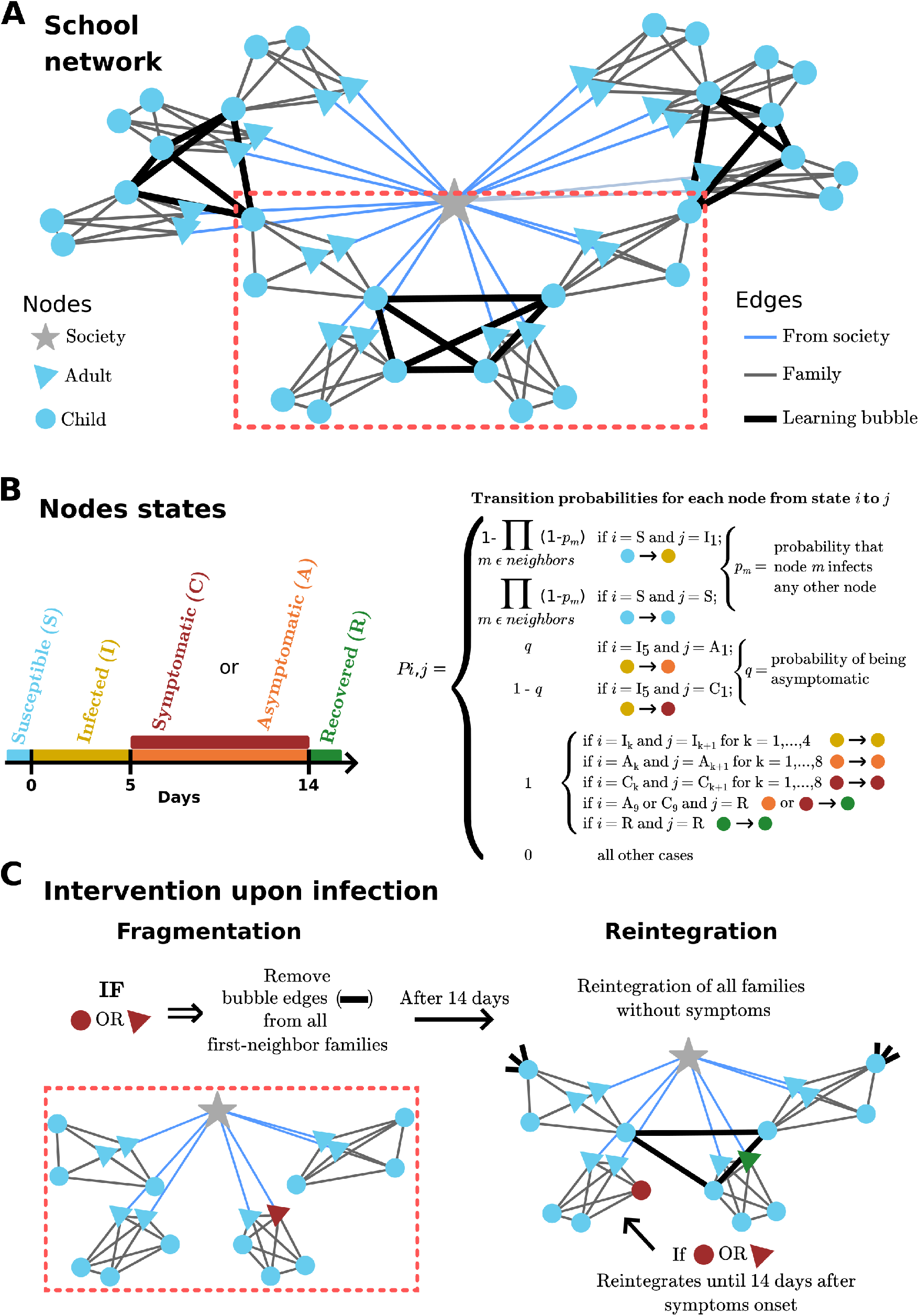
Description of the computational model. **(A)** Representation of a school network with 10 families, where every family is composed by 2 adults (blue triangles) and 3 children (blue circles) nodes. All the members within a family are interacting with no restrictions (gray edges) and only the adults are connected (light blue edge) with the rest of the society (gray star node). Learning bubbles are formed between some of the children nodes (black thick edges). The families that have participating children in a bubble are shown inside the rectangle of red-dotted lines. **(B, Left)** All the nodes in the network (except for the society node) can be in 5 different states: Susceptible (S, light blue), Infected (I, yellow), Symptomatic or Asymptomatic (C or A, red and orange, respectively) and Recovered (R, green). **(B, Right)** The transition from one state (*i*) to another (*j*) depends on the transition-probability matrix (see Materials and Methods for more details). **(C, Left)** When symptoms are detected in a person (adult or child) of the families that participate in a bubble, all bubble edges are removed for 14 days (fragmentation of the bubble). **(C, Right**) After 14 days of fragmentation, the same bubble interactions are reinstated, except those of children whose any family member continue having suggestive symptoms (in that case, the rest of the families return to in-person activities).

## Results

### Analysis of simulated bubbles

We simulated school networks with bubbles of different sizes (*k*=1 [no bubbles], 4, 6, 8, and 20 [regular-size classroom]). In addition, in order to compare this strategy with a school-reopening scenario, we also considered bubbles of size 20 in which a teacher (adult node) is included and all children and the teacher wear facemasks (denoted here and thereafter as 20_mask_). We found the percentage of families with any of its members infected (referred as infected families) after 90 days of simulation increases with bubble size (Fig. 2A). Notably, the school-reopening scenario is similar to a small-bubbles (*k*=4, 6) model, suggesting that allowing normal interactions with children within their bubbles restrict virus propagation in a similar manner to reopening schools with mitigation measures (Fig. 2A). Although the *k*=1 case provides a basal number of infected families through infections from the society node, we wanted to measure directly the frequency in which infections passed through bubble interactions. Therefore, we measured the number of children that got infected through a single infected child of the same bubble (Bubble_R_0_), which is related to the basic reproductive number in epidemiology, but restricted to infections within the bubble (similar to the concept of event R_0_ defined in *23*). We found that for any *k*, Bubble_R_0_=0 in most cases, but the distribution of non-zero Bubble_R_0_ varies with *k* (Fig. 2B and Table S4). In agreement with Fig. 2A, the non-zero Bubble_R_0_ values reveal that some infections do pass to families through the bubbles, and the percentage of breakthrough infections is significantly smaller when the size of the bubble is small (*k*=4) than for *k*=20_mask_ (Fig. 2B). Since a suspected or positive case is detected in a family results in the fragmentation of the bubble or classroom, we also investigated how infections affect the number of days in which children are isolated from their bubbles affecting not only the continuity of the learning process, but also the ability of their parents to go to work. We found that operating in small bubbles (*k*=4-8) is much more effective than reopening schools (*k*=20_mask_ or even if classroom sizes are smaller; Fig. S1) avoiding interruptions of in-person learning (Fig. 2C). In particular, in our 90-day simulations, when *k*=4 most families are isolated less than 20 days, while for *k*=20_mask_ a significant number of families are isolated between 40-50 days, *i*.*e*., about half of the total time (Fig. 2C). We also asked how the number of siblings affects the number of days in which families are isolated and found the number siblings attending bubbles and bubble size proportionally increase the number of days that families need to be isolated (Fig. 2D). Taken together, we conclude that segregating schoolchildren into small bubbles (*k*=4) perform similarly or slightly better than reopening schools with mitigation measures (*k*=20_mask_) in terms of COVID-19 propagation (Fig. 2A, B), and operating in small bubbles is more effective in ensuring longer periods of in-person attendance and reducing the number of missing days of in-person learning with respect to reopening schools (Fig. 2C, D; Fig. S1).

**Fig. 2.**
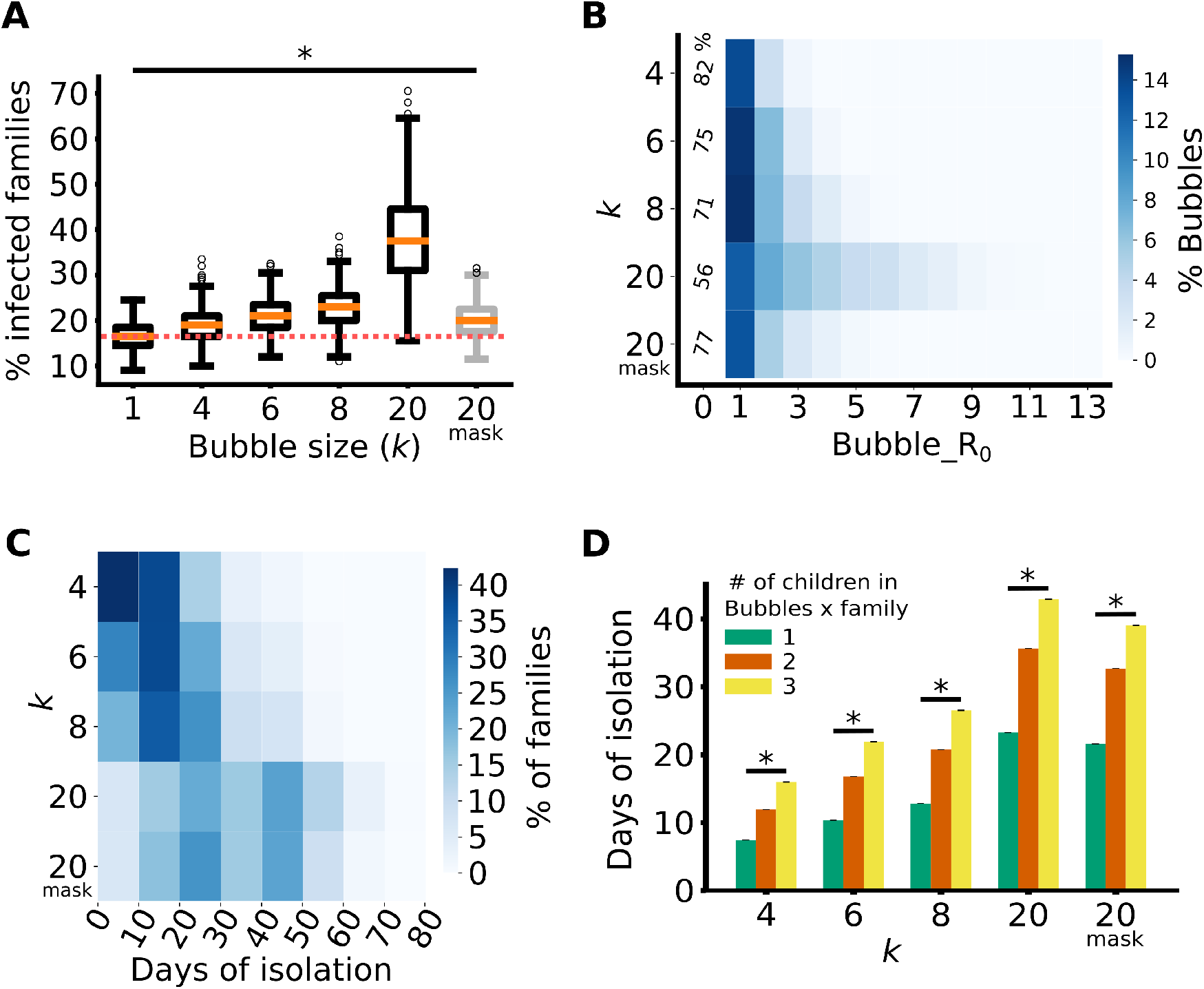
Small learning bubbles have better outcomes than a school-reopening model. (**A**)Average of the percentage of the families in the school network that got infected during the simulations with different bubble sizes (20_mask_ refers to a bubble of size 20 where children wearing mask in the bubbles). The red dotted line marks the median of *k*=1 (no bubbles) for comparison purposes. **(B)** Histogram depicting the Bubble_R_0_ for different bubble sizes, *k*. The percentage of bubbles with an index case are represented by color intensities of a heat map. For better visualization purposes, Bubble_R_0_ = 0 is depicted as a number in the first column. **(C)** Histogram of the number of days (in ranges of 10 days) that families were isolated due to a possible infection for different bubble sizes, *k*. The percentage of families are represented by color intensities of a heat map. (Total number of days in these simulations is equal to 90). **(D)** Number of days that the families were isolated from the bubble, depending on the number of their children that participate in the bubbles program for the different bubble sizes, *k*. (Total number of days is equal to 90). For A and D, we used the Kruskal-Wallis and/or Mann-Withney non-parametric tests as some of the distributions were not normal. ^*^ => p<0.05.

### Robustness of network dynamics to common perturbations

In contrast to computer simulations, real-world implementations do not always occur as planned. For instance, even if masks are enforced in school, in practice, not everyone use them properly or at all times. We wondered how do the number of infected families, Bubble_R_0_, and number of isolation days depend on perturbations in operational variables. For simplicity, we limited our robustness analysis to *k*=4 (small-bubble model) and *k*=20_mask_ (reopening-school model). First, we investigated how the system responds to perturbations in symptoms reporting. In our original simulations, we assumed that parents report symptoms in a daily basis and as soon as suggestive symptoms occur in a family member (*i*.*e*., when in C_1_), bubbles are fragmented the following day. In practice, however, some people may delay reporting of symptoms, simply because sometimes they appear mild at first. We found that when 70% of families report symptoms on time, but 30% report symptoms on the following day (C_1_70 C_2_30), the number of infected families increases in both *k*=4 and *k*=20_mask_ models, but the *k*=4 case appears to be more robust to this perturbation (p-value=3.34 x10^−2^ *vs*. 9.0 × 10^−4^ for *k*=20_mask_; Fig. 3A, left). A similar result is observed in the measurement of Bubble_R_0_ suggesting that this perturbation does not increase significantly infections through the bubbles (Fig. 3A, middle) nor the days that bubbles are fragmented (Fig. 3A, right). Second, we increased the infection rates by 50 or 100% and asked how this perturbation affects the functioning of the bubbles or reopening of schools. The number of infected families proportionally increases with infection rates (Fig. 3B, left) and this is reflected in the distributions of days that bubbles need to be isolated (Fig. 3B, right); however, this does not affect the number of infections through the bubbles (Fig. 3B, middle). This result suggests that infections in the community do not correlate with infections within the bubbles or within classrooms when mitigation measures are in place, in agreement with a recent report (*24*). We also investigated whether exposure to additional adults (teachers using masks) would affect the operation of small bubbles. Our simulations show that when a single teacher or external tutor is added to each bubble of size *k*=4 there is an increasing risk of infection, although this risk can be contained if children in the bubble also use masks (Fig. 3C). This result provides schools willing to segregate children into small bubbles with a choice; they could opt to keep their teachers working remotely and have the children interact without restrictions or have a teacher present but having children to wear masks. Finally, we investigated how robust is to reopen schools when we assume that a certain percentage (25 or 50%) of children not wearing masks (or not wearing them properly). Our simulations show that the percentage of infected families is very sensible to perturbations in mask wearing (Fig. 3D, left), and this increase is likely to infections occurring in school (Fig. 3D, middle). In summary, our analysis suggests that small bubbles (*k*=4) are more robust than reopening schools (*k*=20_mask_) to perturbations in reporting symptoms (Fig. 3A); infections through the bubble are stable in either case when the infections in the society are significantly increased (Fig. 3B); small bubbles are sensitive to adding an external adult, but this can be compensated, if needed, by the use of masks within the bubble (Fig. 3C); and, reopening schools are sensitive to mask wearing (Fig. 3D), confirming that mitigation measures are essential to keep infections controlled (*24*).

**Fig. 3.**
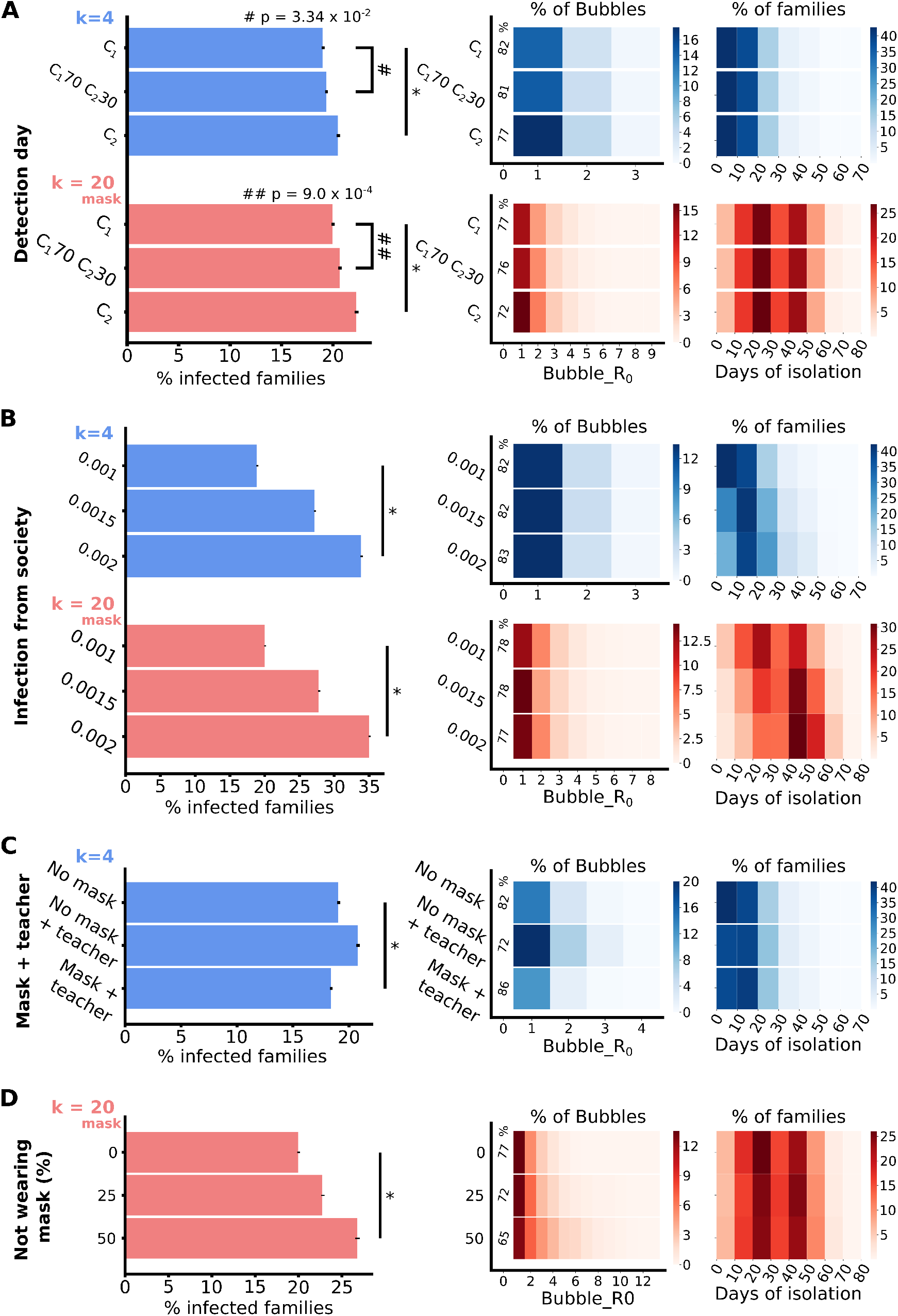
Robustness of the models against common perturbations. **(A-D)** Variations of the percentage of infected families (left), distribution of Bubble_R_0_ (middle), and distribution of isolation days (right) under the following perturbations (blue bars correspond to the small-bubble model [*k*=4] and red bars correspond to the school reopening model [*k*=20_mask_]): **(A)** Fragmentation of a bubble upon a detection of an infected individual is delayed from day 1 (C_1_, top bars) to day 2 (C_2,_ bottom bars) days of symptoms onset, also with 70% detected at C_1_ and 30% at C_2_ (C1_70_C2_30_, middle bars); Increased probability of getting infected from the society (0.001, 0.0015, and 0.002); Including a teacher with and without having children within the bubble using masks (for *k*=4 only); **(D)** Varying the percentage of children not using mask or not using it appropriately (25 and 50%; for *k*=20_mask_ only. For A-D (left panels), we used the Kruskal-Wallis and/or Mann-Withney non-parametric tests as some of the distributions were not normal. ^*^ => p<0.05.

### Learning bubbles data in an elementary school in Mexico City

A small-bubble (*k*=3-5) program was implemented by a private elementary school in Mexico City from October to December of 2020. The program started after 6 months of school closure due to the COVID-19 pandemic, when most parents confirmed that many of their children were already seeing other children and wanted to implement small bubbles in their school (Fig. S2). Participation in the program was voluntary; 50 families (referred as F1-F50) opted-in for the program that consisted of 15 bubbles (referred as B1-B15), some of them with 2 or 3 children in different bubbles (Fig. 4A). Bubbles met alternating weekly in the homes of each of the participants 4 times/week. Children within the bubble interacted without any protection measure. An external teacher or tutor (referred as T1-T13) was included in each of the bubbles at least twice a week; in a few cases the same teacher attended to two bubbles simultaneously (see Fig. 4A). Teachers and parent hosts were told to always use facemasks and obey social distancing when interacting with the children. The operation of the program and collection of data were done entirely by the school, following a specific protocol designed by us for symptoms reporting (Text Box S1-S2) and fragmentation/reintegration of bubbles (Fig. S3-S4). Throughout the duration of the program, each family completed a required daily survey (Text Box S1); in this survey symptoms and other risk factors (such as contact with someone confirmed or highly suspected with COVID-19 disease) were identified. When the criteria of COVID-19 risk (Fig. S3-S4) were met in the daily survey, the affected bubble was fragmented the following day and families in fragmented bubbles were followed up using a different daily survey (Text Box S2). At the beginning of the program, all families were considered susceptible, regardless of whether someone in the family had COVID-19 before the start of the program. The dynamics of how the bubbles behave is shown in Figure 4A. During the first few weeks, infections remained low, correlating with the rate of infections reported by the government of Mexico City (*25*; Fig. 4A). However, following an increase of local cases in the community, some suspected or confirmed cases (marked as Infected in Fig. 4A) started to appear in participating families. During the operation of the program (47 days), 10 families with suspected individuals were detected (we are not sure how many families had confirmed COVID-19 cases since testing was not required by the school; Fig. 4A). We also measured the estimated Bubble_R_0_ in each of the bubbles that were fragmented and found that in 3 of 5 cases, infections did not break through the bubble; and in 2 out of 5 cases, only one additional family may have been infected through the bubble (Estimated Bubble_R_0_=1; Fig. 4B). Notice in contrast to the Bubble_R_0_ obtained from our simulations (Fig. 2B and Materials and Methods) the Estimated Bubble_R_0_ only takes into account symptomatic cases because no testing was required in families of affected bubbles. We also measured the number of days that families were isolated from their bubbles and found that the program was effective in maintaining children learning in their bubbles for the majority of the time (Fig. 4B). Finally, in a post-program survey, the school asked how parents felt while their children attended to their learning bubbles. The majority of the parents responded that they did not feel their family was at risk in this program (Fig. 4D). In addition, from the families that got COVID-19 during the duration of the program, only one family believed that they got infected due to their participation in the program (Fig. 4E). In agreement with our simulations, we conclude that segregating schoolchildren into small bubbles appears to be a safe and effective alternative to reopening schools.

**Fig. 4.**
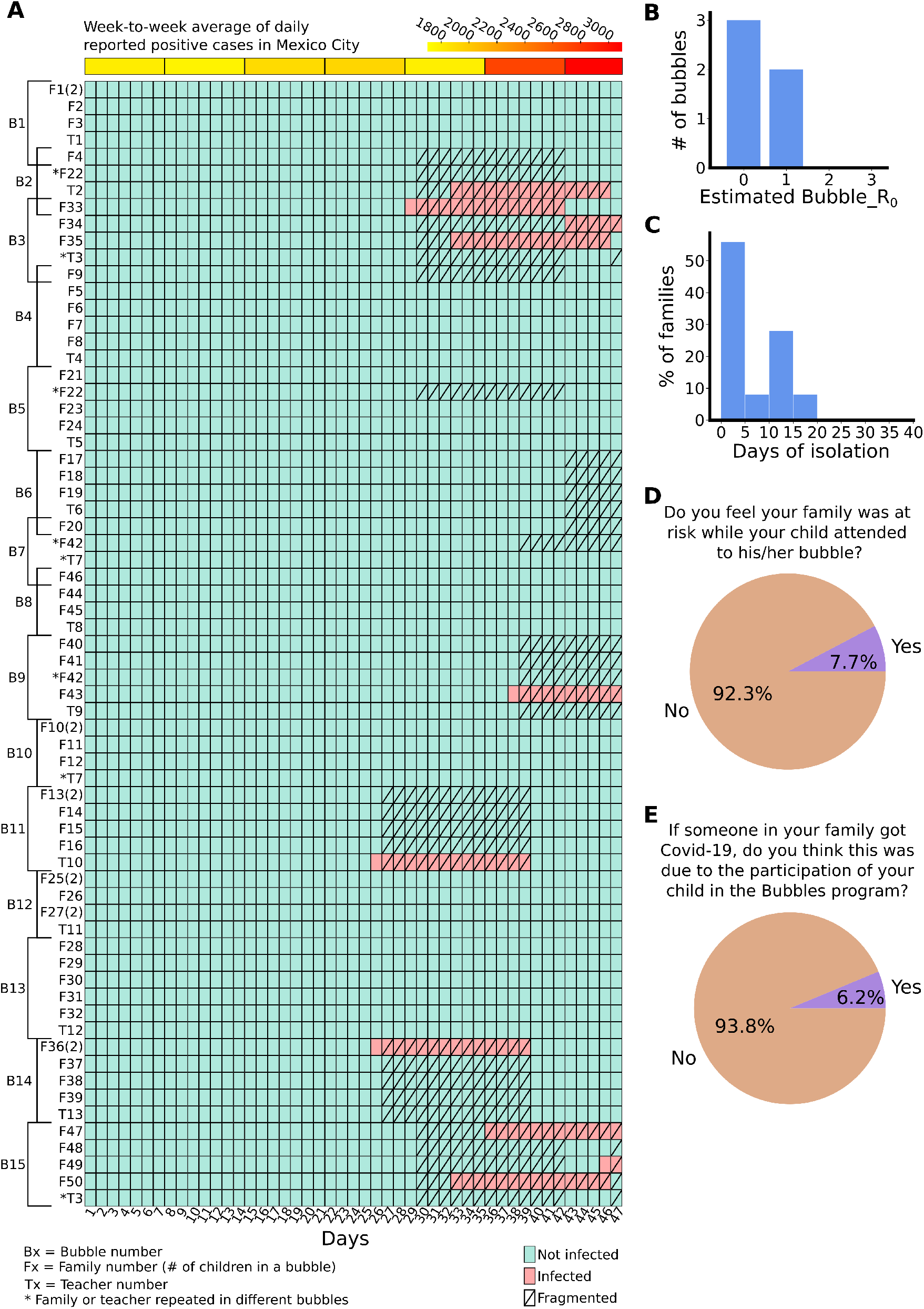
Implementation of a learning bubbles model in an elementary school in Mexico City. **(A)** Dynamics of suspected or confirmed COVID-19 infections in 15 learning bubbles (B1-B15) integrated by children of 50 families (F1-F50) and 13 teachers (T1-T13) from an elementary school in Mexico City. Color boxes represent if any member of a families had COVID-related symptoms (light red) or not (light green) during each of the 47 days that the bubbles operated. At the top, a color map depicts for the weekly average of the daily number of positive cases reported in Mexico City (weekends and holydays not included). **(B)** Histogram of the estimated Bubble_R_0_ within the infected bubbles. **(C)** Histogram of the days that families were forced to interrupt their participation in the bubbles program. **(D, E)** Pie charts showing the opinion of the parents that opted-in for the bubbles program after the 47 days of operation.

## Discussion

Long-term lockdowns experienced during the COVID-19 pandemic are already having a detrimental effect in mental health and learning skills in children (*1, 2, 13–15*), and are causing an increase in family violence, school abandonment, and unemployment (*16*). There is no doubt that most children strongly benefit from in-person learning and ludic activities, but the question that remains is how to do it in a safe and stable manner. While a recent study shows that reopening schools do not correlate with community transmission only when with the appropriate mitigation measures (*24*), implementing these in many schools of developing countries poses financial, logistic, and cultural challenges. We use computational modeling and a real-world implementation to support that segregation of schoolchildren into bubbles while receiving remote instruction from their teachers is an alternative solution. We showed that segregation into small bubbles is not only effective in controlling virus propagation to a similar extent as reopening of schools (Fig. 2A,B; Fig. 3A,B), but also provides additional advantages such as giving children the opportunity to interact without restrictions, it is more stable in maintaining in-person activities when positive cases surge (Fig. 2C), and it is flexible to adding a tutor or teacher if additional mitigation measures (e.g., mask wearing) are included (Fig. 3C). In fact, schools and parents may decide to implement use of facemasks within the bubbles and propagation risks reduce considerably, at the expense that children will not interact without restrictions (Fig. S5). However, the implementation of small bubbles is not free of risks or logistic issues; for example, although with low probability, some children and their families may still get infected through the bubbles (Fig. 4B), it requires management of the daily surveys to assess health checks on families and make decisions on when a bubble needs to be fragmented or reintegrated, it requires parents to be willing to send and receive children in their own homes, and teachers may require the development of additional curricular activities to encourage collaborative and social learning within the bubble.

There are some ways in which our computational model may be varied to take into account specific situations. For example, our simulations in bubbles and classrooms could be improved if testing is conducted in suspected, or even randomly in unsuspected, individuals. We did not consider testing to keep the intervention as affordable as possible for schools and parents, but if testing is incorporated periodically, some asymptomatic individuals may be detected before the virus propagates into the bubbles, reducing the Bubble_R_0_ distributions. In addition, after a fragmentation of a bubble, testing may reduce the number of days in which unaffected children and parents need to be isolated from their in-person activities. Finally, even in communities where reopening of schools is affordable, they may consider operating in small bubbles within their classrooms or offer the formation of external bubbles as an option to some families. This hybrid alternative, if implemented correctly, could be an optimal solution as it reduces virus propagation and days of isolation, and at the same time children are able to return to school buildings.

While our computational model was aimed to explore the effectiveness of segregation of schoolchildren in the current COVID-19 pandemic, it is not limited to this situation. With minimal modifications, the model can be easily adapted to new variants of the virus and other lockdown situations. Furthermore, we believe that working in small bubbles may create new learning alternatives in the post-pandemic world by providing children with the opportunity to gain team-working and collaborative skills.

## Materials and Methods

### The school network and formation of bubbles

Using the NetworkX (https://networkx.org/) python package, we modeled a school network composed of 200 independent families. Each family consisted of 2 adults and 3 children, but not all children were assumed to participate in the bubbles. Each family contributed with at least one child; the second child participated with a probability of 50%, and the third child with a probability of 25%. Bubbles of a specific size *k* were created randomly, only avoiding siblings to be in the same bubble. For each *k*, the code generated 10 bubbles configuration and keeps one with the higher number of bubbles. Children that cannot be accommodated into a bubble were left out. Interactions within a bubble were considered as household interactions, *i*.*e*., with no restrictions, except for when masks are used (e.g., *k*=20_mask_, Fig. 3C, or Fig. S5). Teachers are considered to work remotely (*i*.*e*., not considered in the network) except for *k*=20_mask_ and when explicitly stated in Fig. 3C. When teachers participate in-person, they are considered as solitary adults in the network, connected to the society node. Only one independent teacher is considered per bubble.

All networks begin with all individuals in a susceptible state and dynamics begin when adults begin to get infected from the society node. Each simulation is run for 90 steps (1 step= 1 day) of the model and for each network configuration, we run 500 simulations. The resulting data are stored in a file for further analysis. An example of dynamics of this model for a network of 10 families is illustrated in Movie S1.

### Bubbles fragmentation and reintegration

For every step in the model, the state of each the nodes (except for the society node) within the school network is evaluated. If a node enters a symptomatic state C_1_, the corresponding family is considered possibly infected and all the bubbles in which they participate are fragmented (edges in the network removed) for at least 14 days or until no member of the family remains symptomatic. When a family of a fragmented bubble that is not the infected family has additional children in other bubbles, these children are separated from their corresponding bubble, but the bubble is not fragmented. After 14 days, bubble edges are reinstated, except for the families that continue to have symptomatic individuals.

### Nodes infection from their neighbors or from the society

Every node in the network (children and adults) may be infected from their first neighbors nodes with a probability given by the transition matrix depicted in Figure 1B. This matrix considers the probability (*p*_m_) for each node is infected by its m-th neighbor which are obtained taking into account the type of interaction and whether the neighbor is symptomatic or asymptomatic (Table S1) and the day of contact (Table S2) to have a resultant individual probability (Table S3). If the node is an adult, one of the neighbors is the society node which probability is given by the rate of virus propagation in the community (see first row in Table 2 and Fig. 3B). Assuming independency of events, the probability of a node of becoming infected (I_1_) at the following step is given by.

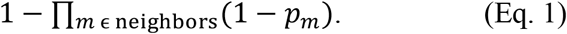

### Infection through a bubble

To compute the Bubble_R_0_ in our simulations (Fig. 2–3), we consider the case when a child gets infected from a family member (index case) and count all the bubble neighbors that get infected by this event (including secondary transmissions by other children in the bubble that got infected by the index case). This integer number is what we define as Bubble_R_0_.

In practical implementations (Fig. 4), however, when there are multiple sources of infection (some of them unknown as they may be asymptomatic, non-tested individuals), it is impossible to know for sure if a person was infected from a specific individual. In this case, we can obtain a crude estimate of Bubble_R_0_ (Estimated Bubble_R_0_; Fig. 4B) as follows. Given a symptomatic individual (child or adult), we considered infections in families that participate in bubbles with family members of the symptomatic individual for the duration of bubble fragmentation (14 days). The Estimated Bubble_R_0_ is given by the number of families with at least one symptomatic or confirmed individual during these 14 days (excluding the index-case family). A comparison of numerical Bubble_R_0_ and Estimated Bubble_R_0_ values for different values of *k* are shown in Table S4.

### Data analysis and visualization

All the data obtained from the simulations was analyzed using the Python3 programming language (https://www.python.org/). The packages used for the analysis were Pandas (https://pandas.pydata.org/) and Numpy (https://numpy.org/), for the statistical test we used the Scipy (https://www.scipy.org/) package and the visualization was done with the Matplotlib (https://matplotlib.org/) package. Statistical tests used in study are indicated in the corresponding figure legends.

### Bubbles program implementation

The population of study was composed of children from a private elementary school (K-6) located in Mexico City. The participation on the program was completely voluntary (a fully online program was available in parallel for families that were not interested or were considered at high risk for the bubbles program). Parents or tutors of the children agreed to share non-personal data including the appearance and evolution of COVID-19-associated symptoms in their families for the duration of the program with the purpose of data analysis and publication. Data was analyzed retrospectively and authors did not participate in its implementation. The identity of the participants was unknown to the authors at all times.

Bubbles were formed based on the following criteria: (1) 3-5 children per bubble; (2) when siblings were in the same grade, they were put in the same bubble; (3) one teacher or tutor was assigned to each bubble and attended in-person activities with the children at least twice a week. Only in a few cases (marked with a * in Fig. 4A) a teacher participated in two bubbles. Decisions on fragmentation and reintegration of bubbles were taken based on the answers that the parents provided in daily surveys (Text Box 1-2) using the criteria outlined in Fig. S3-S4.

At the time of the start of the program none of the parents or teachers were vaccinated; it is unknown which individuals had gotten the COVID-19 disease before the program started.

## Data Availability

All data are available in the main text or the supplementary material.

## Acknowledgments

We thank the following individuals for discussions and contributions through the development of this project: Jacobo Tronllan, Leobardo Emmanuel Soria, Jack Zagha, Norma Ayala, Ruth Franco, Graciela Galante, Vicky Halabe, Esther Laniado, Lucía Dominguez, Renán Escalante, Marycruz Flores, Abraham Mustri, and Eduardo Ochoa.

## Author contributions (CREDiT format)

Conceptualization: LMMN, MN

Methodology: LMMN, MN

Investigation: LMMN, MN

Visualization: LMMN

Project administration: MN

Supervision: MN

Writing – original draft: MN

Writing – review & editing: LMMN, MN

