## Supplementary Material for "Segregation of children into small groups for in-person learning during the COVID-19 pandemic"

### Supplementary Text

#### Text Box S1

**Daily Survey for Families in the Bubbles Program**

1. Please perform a simple smell and taste test for every person in your household. For the smell test, smell inside a jar of coffee and make sure you clearly distinguish the smell. For the taste test, try something sweet. Does anyone in your household have an absence or diminished smelling or tasting sense?    YES    NO

2. Use a regular thermometer to measure the body temperature for every person in your household. Does anyone has a temperature higher than 37.5°C?  YES    NO

3. Please indicate if someone in the family present any of the following symptoms: Dry cough, Fever, Unusual fatigue, Difficulty for breathing, Sore throat, Diarrhea, Runny nose, Nausea and/or vomiting, Muscular or articular ache, Headache, Chest pain, Irritability in children below 5 years of age, Shivers, Diminished smelling or tasting sense, Conjunctivitis.

4. Does anyone in your household was in contact with someone suspected or confirmed for COVID-19?    YES     NO

5. Does anyone in your household returned today from a flight?   YES    NO

6. Does anyone in your household attended to a social meeting or party with more than 30 people?  YES   NO  (If yes, please describe)

7. (Only on Sundays) Did your children meet with other children of their bubble during the weekend?   YES    NO

#### Text Box S2

**Daily Survey for Families in a Fragmented Bubble**

1. Please perform a simple smell and taste test for every person in your household. For the smell test, smell inside a jar of coffee and make sure you clearly distinguish the smell. For the taste test, try something sweet. Does anyone in your household have an absence or diminished smelling or tasting sense?    YES    NO

2. Use a regular thermometer to measure the body temperature for every person in your household. Does anyone has a temperature higher than 37.5°C?  YES    NO

3. Do people in your household present new symptoms today? (If yes, please describe)

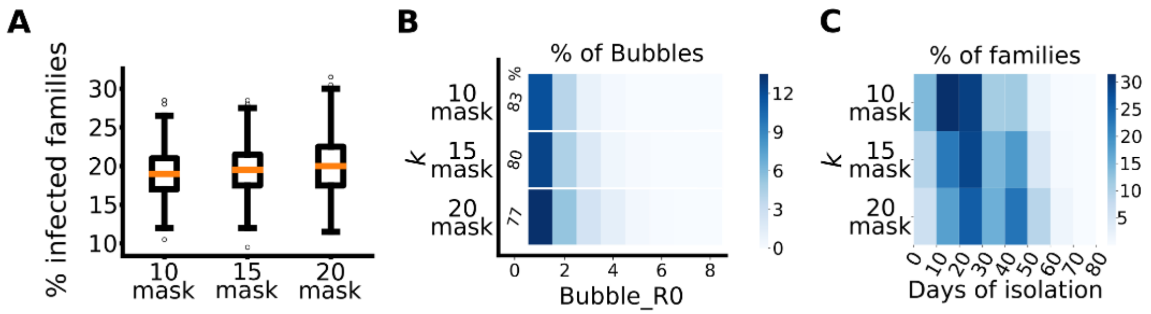


**Fig. S1. Reducing the size of a classroom (comparison of *k* = 10, 15, and 20 + wearing masks) improve the outcomes of the model. (Analysis was done as in Fig. 2A-C; see legend for details).**

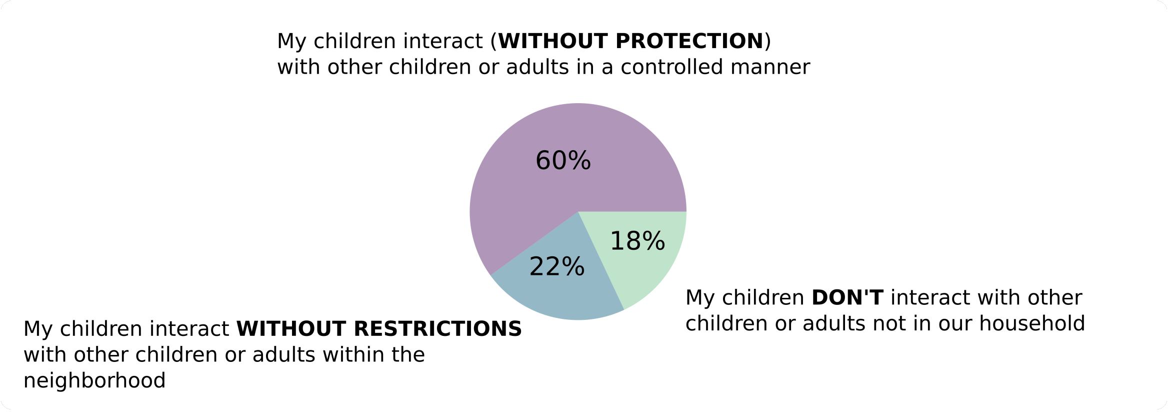


**Fig. S2. Survey applied to all parents in the elementary school in July of 2020 that motivated the implementation of the bubbles program.**

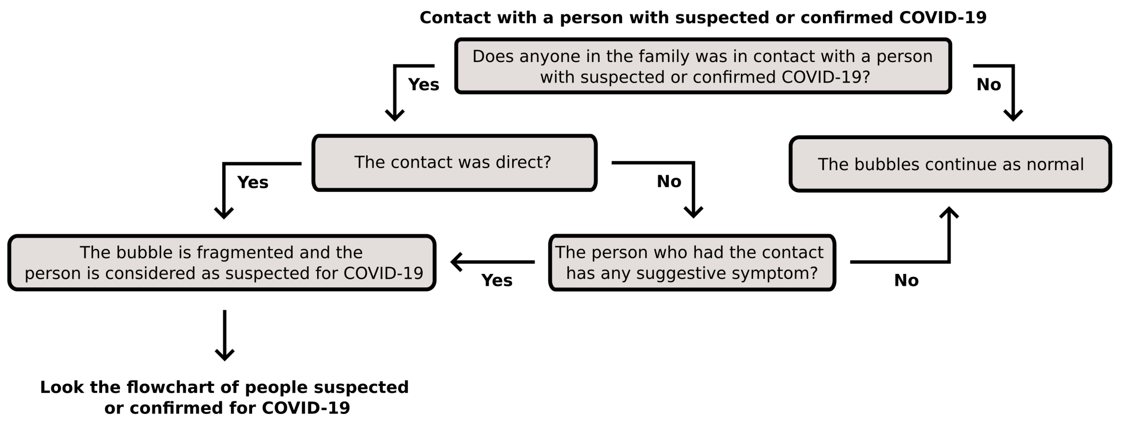


**Fig. S3. Flowchart of the actions followed by the school when someone was a contact of a person suspected or confirmed for COVID-19.**

This is the protocol followed by school administrators when parents report a contact that is strongly suspected of COVID-19 in the daily survey.

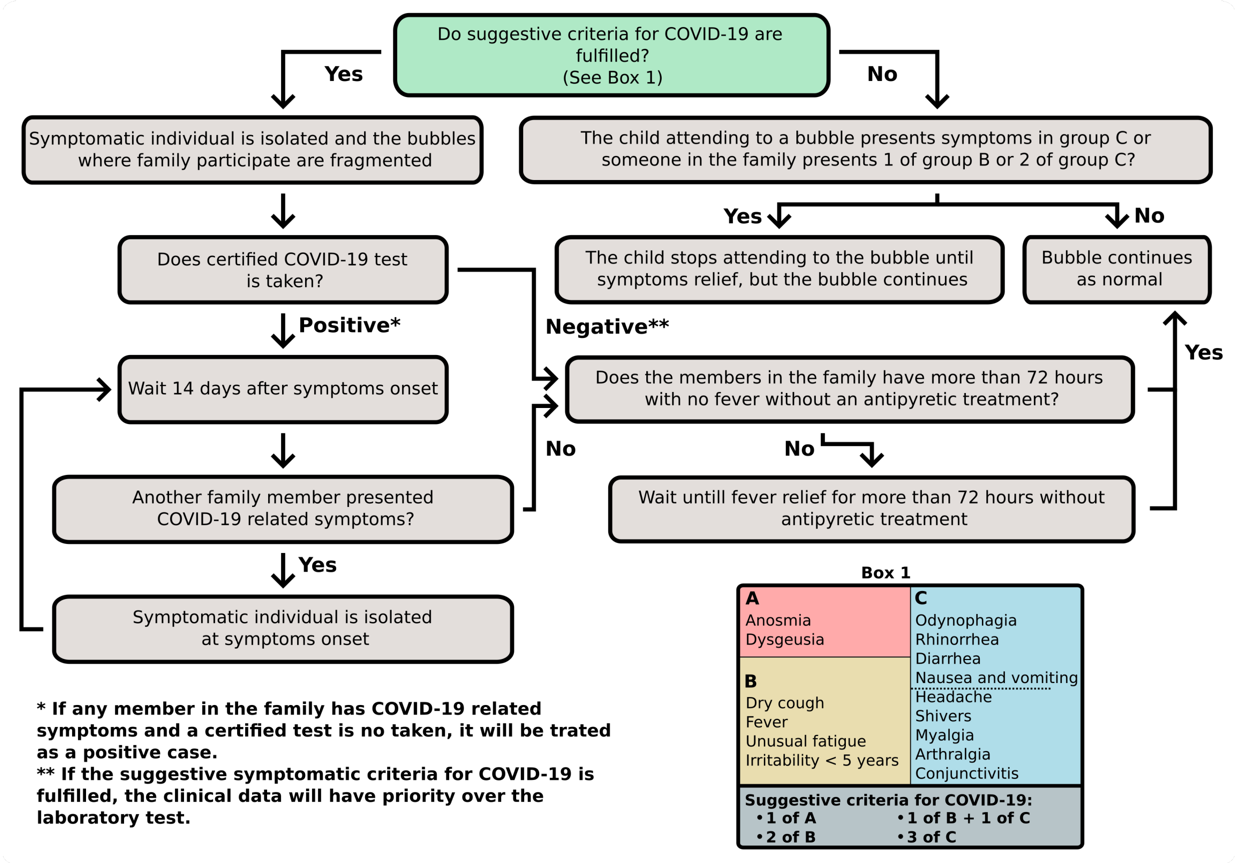


**Fig. S4. Flowchart of the actions followed by the school to determine whether symptoms reported in the daily survey suggest a COVID-19 case even when no testing is available.**

This is the protocol followed by school administrators when parents report symptoms in the daily survey.

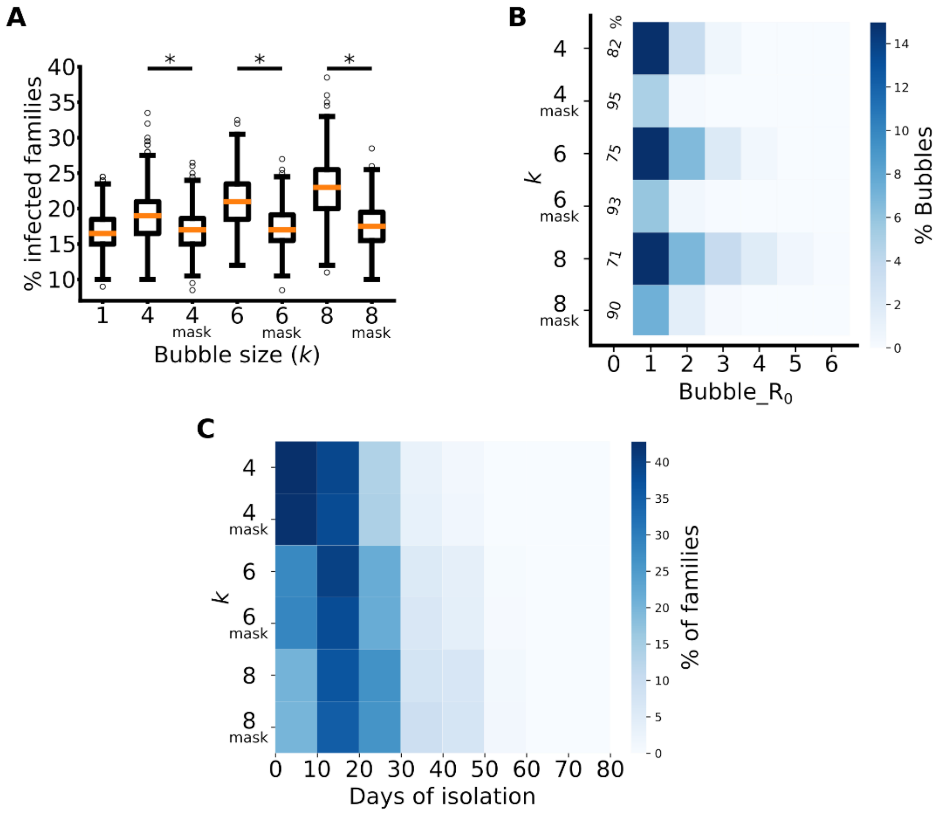


**Fig. S5. Using masks in small bubbles improves COVID-19 transmission outcomes. (comparison of *k* = 4, 6, and 8 with and without masks), but schools may consider the importance of unrestricted children interactions. (Analysis was done as in Fig. 2A-C; see legend for details).**

**Table S1. Parameters obtained from the literature (SARS-CoV2 variant α) that were taken into account in the simulations.**

| Symbol | Description of Probabilities | Numeric value | Bibliographic Reference |
|---|---|---|---|
| Soc_A | Society → Adult | 0.001<br>(High transmission rate) | *26* |
| Sym_A_S | Adult → Spouse | 0.378 | *27* |
| Sym_A_A | Adult → Other Adult | 0.283 | *27* |
| Sym_A_C | Adult → Child | 0.168 | *27* |
| Sym_C_A | Child → Adult | A_A * 0.63 | *28* |
| Sym_C_C | Child → Child | A_C * 0.63 | *28* |
| Asym | Multiplicative protection factor when contact is an asymptomatic individual | Probability * 0.58 | *29* |
| Mask | Multiplicative protection factor when using a mask | Probability * 0.21 | *30* |
| To_be_asym | Probability of being asymptomatic | Adult = 0.17<br>Child = 0.35 | *29* |

**Table S2. Distribution of probabilities depending on the day of contact. Values taken from ref. (*31*).**

| Days (X) after symptoms onset ($D_x$) | Nodes State | Relative probability |
|---|---|---|
| $D_{-5}$ | $I_1$ | 0.071 |
| $D_{-4}$ | $I_2$ | 0.214 |
| $D_{-3}$ | $I_3$ | 0.642 |
| $D_{-2}$ | $I_4$ | 0.785 |
| $D_{-1}$ | $I_5$ | 0.928 |
| $D_0$ | 0 | 1 (symptoms onset) |
| $D_1$ | $A_2$ or $C_2$ | 0.928 |
| $D_2$ | $A_3$ or $C_3$ | 0.785 |
| $D_3$ | $A_4$ or $C_4$ | 0.642 |
| $D_4$ | $A_5$ or $C_5$ | 0.357 |
| $D_5$ | $A_6$ or $C_6$ | 0.214 |
| $D_6$ | $A_7$ or $C_7$ | 0.143 |
| $D_7$ | $A_8$ or $C_8$ | 0.071 |
| $D_8$ | $A_9$ or $C_9$ | 0.035 |

**Table S3. Individual probabilities $p_i$ that appear in the transition matrix (Fig. 1B).**
Actual values are computed using the parameters of Tables S1 and S2.

| **Type of contact** | **Individual probability to infect ($p_m$) assuming that m is a neighbor** |
|---|---|
| Symp_Adult - Spouse | Sym_A_S*$D_x$ |
| Symp_Adult - Child | Sym_A_C*$D_x$ |
| Symp_Child - Adult | Sym_C_A*$D_x$ |
| Symp_Child - Child | Sym_C_C*$D_x$ |
| Asymp_Adult - Spouse | Asym*Sym_A_S*$D_x$ |
| Asymp_Adult - Child | Asym* Sym_A_C*$D_x$ |
| Asymp_Child - Adult | Asym* Sym_C_A*$D_x$ |
| Asymp_Child - Child | Asym* Sym_C_C*$D_x$ |

**Table S4. Numerical values of the percentage (%) of bubbles that correspond to different Bubble_$R_0$ (as in Fig. 2B) and Estimated (E) Bubble_$R_0$ for the different *k***

| | **Bubble_$R_0$ or Estimated (E) Bubble_$R_0$** | | | | | | | | |
|---|---|---|---|---|---|---|---|---|---|
| ***k*** | 0 | 1 | 2 | 3 | 4 | 5 | 6 | 7 | 8 |
| 4 | 82.18 | 13.86 | 3.39 | 0.56 | 0.00 | 0.00 | 0.00 | 0.00 | 0.00 |
| 4 (E) | 83.62 | 13.96 | 2.08 | 0.34 | 0.00 | 0.00 | 0.00 | 0.00 | 0.00 |
| 6 (R) | 75.51 | 15.15 | 6.79 | 2.04 | 0.48 | 0.04 | 0.00 | 0.00 | 0.00 |
| 6 (E) | 73.73 | 19.78 | 4.70 | 1.36 | 0.35 | 0.09 | 0.00 | 0.00 | 0.00 |
| 8 (R) | 71.16 | 15.46 | 7.10 | 3.81 | 1.83 | 0.56 | 0.07 | 0.00 | 0.00 |
| 8 (E) | 65.70 | 23.87 | 6.45 | 2.39 | 0.94 | 0.48 | 0.15 | 0.01 | 0.00 |
| 20 (R) | 56.46 | 12.82 | 8.00 | 6.18 | 4.93 | 3.69 | 3.28 | 2.10 | 1.44 |
| 20 (E) | 30.13 | 28.74 | 16.59 | 9.50 | 5.08 | 3.11 | 1.95 | 1.25 | 1.03 |
| $20_{mask}$ (R) | 77.14 | 13.32 | 5.29 | 2.47 | 1.23 | 0.41 | 0.09 | 0.04 | 0.01 |
| $20_{mask}$ (E) | 45.05 | 32.87 | 14.71 | 4.48 | 1.68 | 0.78 | 0.24 | 0.13 | 0.04 |

**Movie S1.**

Example of the dynamics of a school network using 10 families in one simulation of 66 days (steps) assuming *k*=4, a very high (0.002) probability of infection from the society (*26*), and symptoms detection day at $C_1$. The nodes, edges, and colors are as described in Fig. 1.

**Data S1. (Spreadsheet_for_fig2_and_fig_S5)**

Numerical values corresponding to the color-map values in Fig. 2B, Fig. 2C, and Fig. S5.

**Data S2. (Spreadsheet_for_fig3_and_fig_S1)**

Numerical values corresponding to the color-map values in Fig. 3A-D and Fig. S1.

**Separate files names:**

Spreadsheet_for_fig2_and_fig_S5
Spreadsheet_for_fig3_and_fig_S1

**Link for the code:**

https://github.com/aracar9/Bubbles.git
